# Novel Digital Gastric Alimetry® Symptom Scales for Use in Gastroduodenal Disorders and Validation against Rome IV Criteria

**DOI:** 10.1101/2025.02.19.25322571

**Authors:** Armen A. Gharibans, I-Hsuan Huang, Chris Varghese, Gabriel Schamberg, Sarvnaz Taherian, Nicky Dachs, Mikaela Law, Stefan Calder, Christopher N. Andrews, Jan Tack, Greg O’Grady

## Abstract

**Background:** Patients with chronic gastroduodenal disorders present with overlapping symptoms. Guidelines emphasize symptom-based criteria, but clinical evaluations are inconsistent due to non-standardized assessments and recall bias. Gastric Alimetry® is a non-invasive test of gastric function enabling real-time symptom evaluation via a standardized app.

**Methods:** Participants meeting Rome IV criteria for functional dyspepsia (FD) and/or chronic nausea and vomiting syndrome (CNVS) underwent a Gastric Alimetry test, including a meal challenge, with symptoms recorded every 15 minutes in the app. Based on time-of-test symptoms, four novel scores were developed: nausea/vomiting, postprandial distress, epigastric pain, and burning/reflux. Group differences were analyzed using pairwise t-tests, and Rome IV classifications were predicted via logistic regression. Remote moderated usability testing assessed score acceptability.

**Key Results:** Among 109 participants (79% female, 18-80 yrs), 54 met criteria for CNVS with/without FD, 41 for postprandial distress syndrome (PDS) only, and 14 for epigastric pain syndrome (EPS) with/without PDS. Symptom scores aligned with Rome IV classifications (*p*<.05 for CNVS and EPS). Logistic regression showed good discrimination for CNVS (AUC=0.85) and EPS (AUC=0.80), and moderate discrimination for PDS (AUC=0.68). Usability testing confirmed clinical utility and ease of use.

**Conclusions & Inferences:** Gastric Alimetry symptom scores align with Rome IV classifications, with real-time patient-reported snapshots accurately reflecting chronic symptom burden. These scores provide a clinically applicable diagnostic tool alongside simultaneous physiological gastric function assessments.

**Key Points:**

- Four novel Gastric Alimetry symptom scores summarize the relative severity of symptoms in subgroups aligned with Rome IV classifications.
- The proposed time-of-test symptom scores showed moderate-to-good ability to predict diagnoses made using the Rome IV criteria.
- Usability testing with eight clinicians showed that the scores provided an easy-to-use and clinically useful tool to complement diagnosis of gastroduodenal disorders.

## Introduction

Diagnosing and managing chronic gastroduodenal disorders, including functional dyspepsia (FD) and chronic nausea and vomiting syndrome (CNVS), is challenging due to overlapping symptoms.^1,2^ While gastric emptying, the current gold-standard for diagnosis, is informative, it does not reliably distinguish between these conditions, which require different management approaches.^3,4^

Current diagnostic guidelines emphasize self-reported symptom profiling to guide diagnosis,^5–7^ but such assessments are frequently limited by non-standardized clinical methods and recall bias. Additionally, clinicians often lack time to perform detailed symptom evaluations with validated questionnaires, and patients may struggle to differentiate specific gastrointestinal symptoms and their relationships to meals.^8^ Consequently, a simple and reliable clinical tool for accurate symptom profiling is highly desirable.

Gastric Alimetry^®^ (Alimetry, New Zealand) is a new, non-invasive test of gastric function that enables simultaneous body surface gastric mapping (BSGM) and standardized symptom evaluation via the validated Gastric Alimetry App.^9,10^ The App incorporates symptom descriptions alongside pictograms shown to improve accuracy in patient symptom classifications.^8,11^ The App, in conjunction with a 4.5-hour test protocol that includes a meal challenge, has demonstrated robust validity and ease of use.^12^ This App therefore presents a convenient and reliable option for symptom profiling in clinical practice.

This study aimed to leverage the outputs from the Gastric Alimetry App to generate a novel set of symptom scores, providing a concise summary of key symptom profiles that align with current gastroduodenal disorder classifications.

## Materials & Methods

Data were collected as part of a prospective observational cohort study conducted in Leuven, Belgium (ethical approval from the Ethics Committee Research UZ Leuven S65541). Participants meeting Rome IV criteria for FD and/or CNVS were included.

Patients underwent a standard 4.5-hour Gastric Alimetry test, comprising a 30-minute fasted baseline, consumption of a ~250 kcal meal, and 4-hr post-prandial recording. The Gastric Alimetry App, which includes a gastroduodenal symptom pictogram set validated in both adults and children, was used (**Figure 1A**).^11,12^ Throughout the test, patients used a dashboard interface to record their symptoms, both at will and in response to 15-minute notifications. Ten symptoms were captured: six continuous symptoms using 0 (none) to 10 (most severe imaginable) Likert scales (nausea, upper abdominal pain, bloating, heartburn, stomach burn, and excessive fullness); three event logs (vomiting, belching, and reflux); and one score from 0 to 10 entered immediately postprandially (early satiation). The App has demonstrated 100% patient compliance.^12^

**Figure 1.**
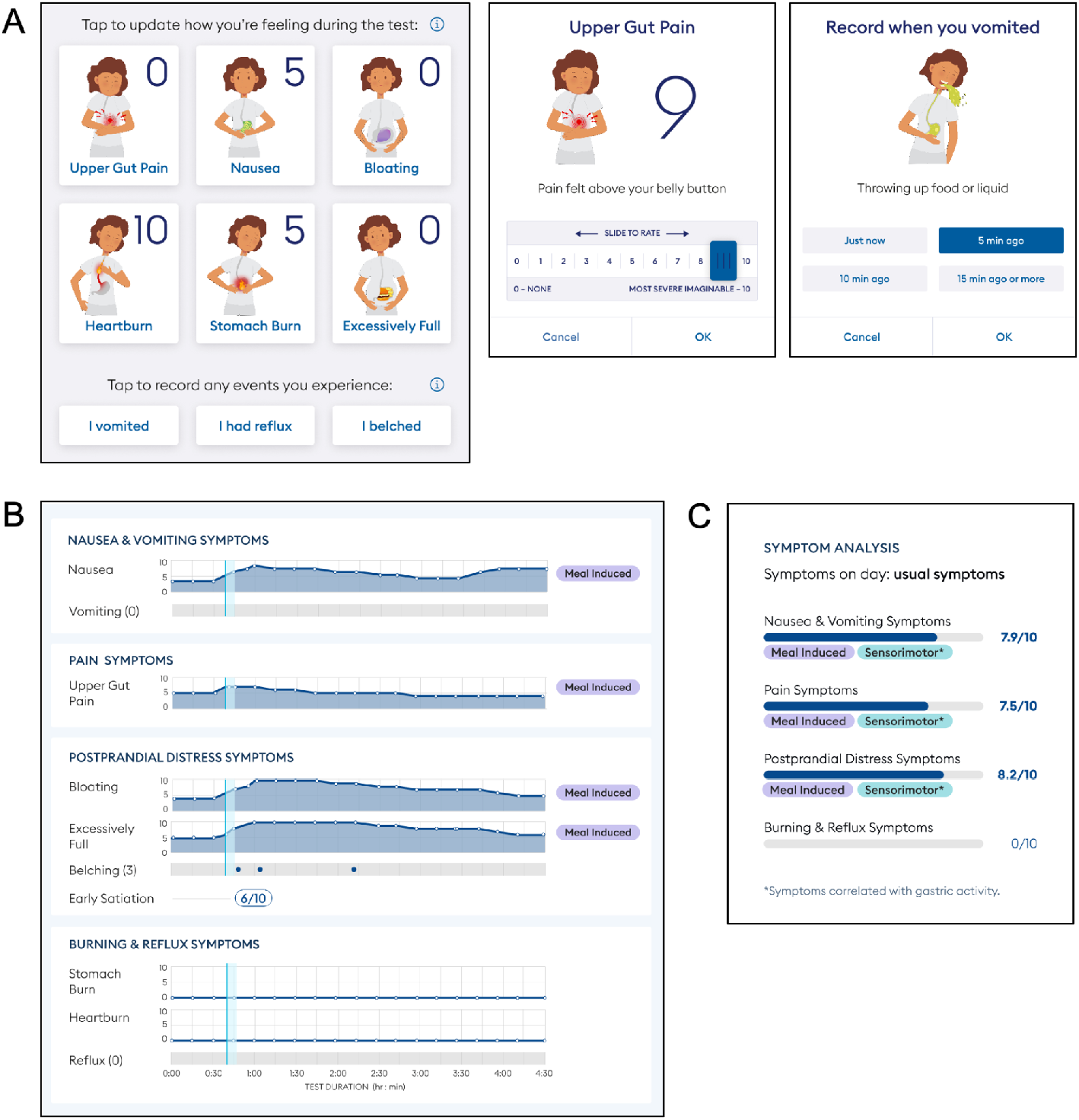
**A)** Patient symptom dashboard in the Gastric Alimetry App, showing six continuous symptoms and three symptom events. Symptoms are recorded over the course of a test meal (30 min baseline; 4 hr postprandially). Update notifications are provided every 15 min and patients can update symptoms or events at any time using the dashboard display. Examples are shown for one continuous symptom example (upper gut pain) and one symptom event (vomiting). **B)** Clinician output, displaying the time-of-test symptom curves and symptom event markers, structured within the four novel score domains. Symptom descriptors (e.g. ‘Meal Induced’) are provided as ‘tags’. **C)** Summary symptom score display, generated from the data in (B), including symptom tags.

Four novel symptom scores (0-10 scale) were developed to summarize nausea/vomiting, postprandial distress, epigastric pain, and burning/reflux, using the calculations described in **Supplementary Text S1**.

A clinical report interface was developed to summarize captured symptom results graphically (**Figure 1B**), along with a summary symptom report (**Figure 1C**). Descriptive ‘tags’ were automatically generated to indicate whether symptoms exhibited a meal-induced, meal-alleviated, late-onset, or continuous profile, and/or a sensorimotor profile (defined as correlation between gastric amplitude and symptoms) (**Figures 1B,C**).^13^ These tags, previously proposed based on rule-based criteria (**Supplementary Text S2**), have demonstrated utility in further subclassifying the origin and profile of gastroduodenal symptoms in relation to a meal.^13–15^

Participants were also classified into standard ROME IV diagnostic categories: 1) CNVS with/without FD, 2) postprandial distress syndrome (PDS) only, and 3) epigastric pain syndrome (EPS) with/without PDS.^1^

Group differences were analyzed using pairwise t-tests. Logistic regression was used to predict Rome IV classifications based on the novel symptom scores, assessing the alignment of these scores with Rome IV classifications and their ability to reflect chronic symptom burden.

Qualitative feedback on the usability and acceptability of the symptom scores was obtained through remote, moderated ‘think-aloud’ usability tests with clinicians. Clinicians were shown two independent reports and asked to interpret them as they would with their own patients. During these sessions, clinicians were encouraged to verbalize their interpretation process, while the moderator explored areas of confusion or difficulty. Sessions were recorded, transcribed, and assessed via thematic analysis.

## Results

A total of 109 participants were recruited (n = 86, 79% female; age range 18-80 years; BMI 16.2-36.9 kg/m^2^), comprising 54 meeting criteria for CNVS with/without FD, 41 for PDS only, and 14 for EPS with/without PDS.

Participants interacted with the App on average every 14.3 minutes (SD = 4.5), reflecting a high level of engagement. Additionally, the average interval between symptom logs for any individual patient did not exceed 18.7 minutes, confirming complete compliance.

Pairwise analysis revealed strong alignment between the novel Gastric Alimetry symptom and Rome IV classifications, effectively discriminating between Rome IV diagnostic groups (**Figure 2A**). The CNVS group exhibited a significantly higher nausea/vomiting score than the PDS group (p = .004). The PDS group’s highest score was for postprandial distress. The EPS group demonstrated significantly higher pain scores compared to both the CNVS (p = .002) and PDS (p < .001) groups, as well as elevated burning/reflux scores compared to the PDS group (p = .033).

**Figure 2.**
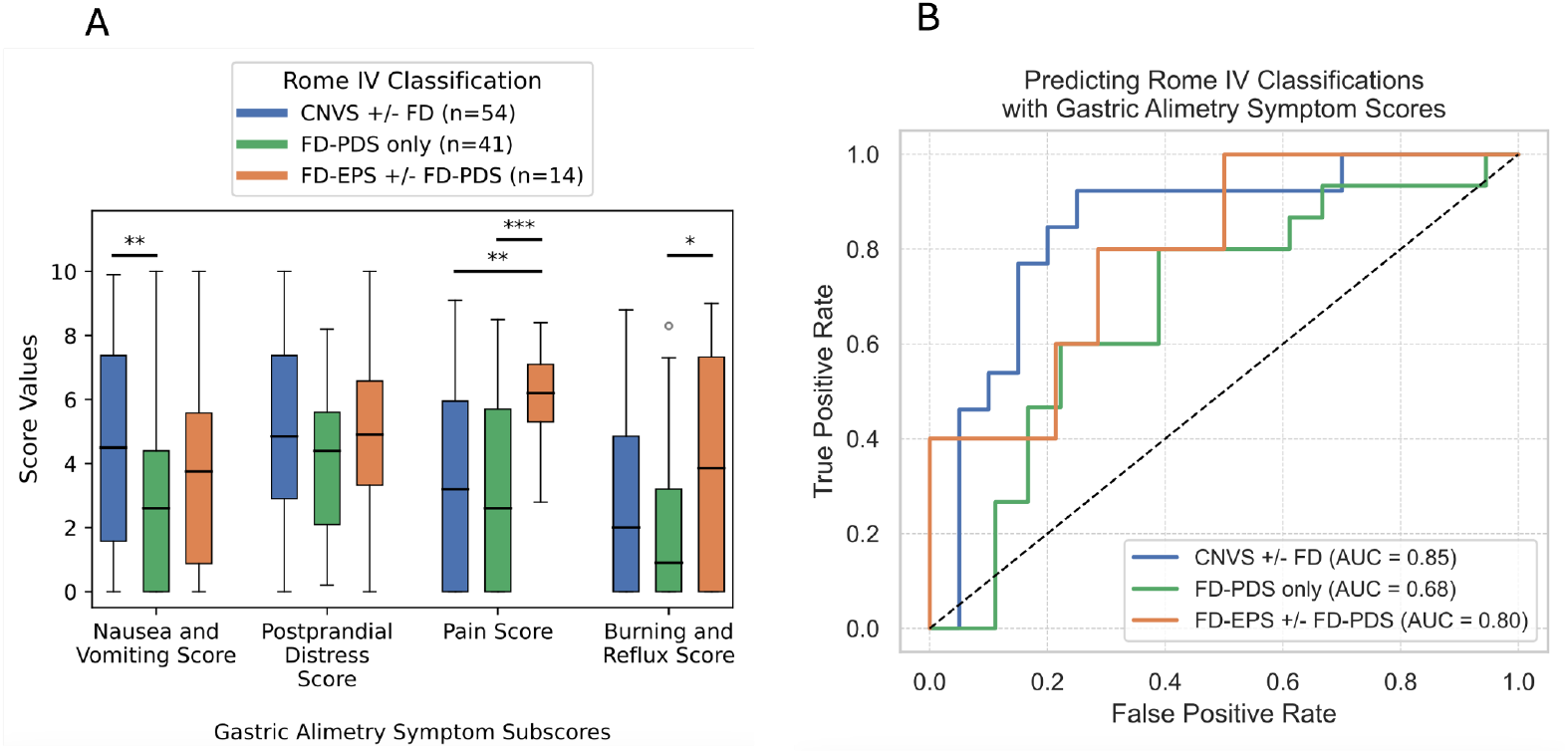
**A)** Box plots displaying the distribution of Gastric Alimetry symptom scores for nausea/vomiting, postprandial distress, pain, and burning/reflux across participants classified by Rome IV criteria (Chronic Nausea and Vomiting Syndrome (CNVS) with/without Functional Dyspepsia (FD), FD with postprandial distress syndrome (PDS) only, and FD with epigastric pain syndrome (EPS) with/without PDS) (*p<.05, **p<.01, ***p<.001). **B)** ROC curves showing the performance of a logistic regression model in discriminating between Rome IV diagnostic groups based on Gastric Alimetry symptom scores. The diagonal dashed line represents the reference for no discrimination (AUC = 0.50).

Logistic regression analysis demonstrated good discrimination for CNVS (AUC = 0.85) and EPS (AUC = 0.80), and moderate discrimination for PDS (AUC = 0.68) based on the four symptom scores (**Figure 2B**). Model details are provided in **Supplementary Text S3**.

Eight clinicians with a range of experience levels using Gastric Alimetry in clinical and/or research settings participated in usability testing. All clinicians agreed on the usefulness and acceptability of the symptom scores. They highlighted that the scores followed existing clinical guidelines, matching their mental models for diagnosis. The scores enabled clinicians to objectively and rapidly compare symptom clusters to identify which were dominant.

## Discussion

This study introduced a set of novel symptom scores, derived from a standardized digital symptom assessment with meal challenge, and validated them against the Rome IV criteria in a cohort of patients with FD and CNVS. While Rome IV categories represent a chronic symptom burden and exhibit significant overlap,^13^ our findings demonstrate that the novel scores effectively differentiate Rome IV diagnostic groups based on a single meal assessment. Together with the novel clinical interfaces presented in **Figure 1**, which were found to be user-friendly and clinically useful, these scores offer a valuable new diagnostic aid for symptom profiling.

Both the Rome criteria and current clinical guidelines emphasize categorizing patients into nausea/vomiting-predominant, PDS-predominant, pain-predominant, or heartburn/reflux categories to appropriately direct management.^5–7^ However, obtaining accurate and detailed symptom insights within time-limited clinical settings can be challenging. The App-based profiling platform described here offers a practical alternative or proxy estimate for the Rome criteria, utilizing pictograms, a digital interface, and summary scores that are all now comprehensively validated.^11,12^ Real-time symptom evaluation may also offer advantages over traditional retrospective questionnaire-based assessments by overcoming recall and mood-congruent biases.^16,17^ In addition, the use of a standardized meal provides a uniform challenge to the upper gastrointestinal tract and eliminates any influence of self-imposed dietary restrictions on daily symptom patterns and severities.^18^

A key strength of the current symptom profiling platform is its ability to deliver a clinically actionable report of patient symptoms in real time alongside physiological assessments. Usability testing confirmed the report’s ease of use and its significant contribution to clinical assessment and diagnosis. This system is currently integrated into the Gastric Alimetry BSGM platform for motility assessment;^9,10^ and the system could also be applied alongside other diagnostic tests, such as gastric emptying testing.^15^ Robustly profiling symptoms during physiological testing is a key method for defining pathophysiological associations and causality in gastrointestinal motility disorders, and we therefore anticipate widespread adoption of this platform in future research.^13,19–21^ In addition, the Gastric Alimetry App optionally includes a validated scale that assesses mental health, including subscales for depression, stress, and anxiety specifically tailored to gastroduodenal disorders, providing additional multimodal inputs for comprehensive profiling of these multifaceted disorders.^22^

A limitation of the study is the use of a single cohort from one country (Belgium). Further validation in diverse patient populations would be valuable. Nevertheless, given the demonstrated global consistency of Rome IV classifications, the results presented here are likely to be generalizable to other populations.^23^

In conclusion, this paper introduces novel digital symptom summary scores for assessing patients with gastroduodenal disorders. The scores are validated, easy to use, and align with Rome IV classifications. They provide a practical diagnostic aid that will be particularly useful alongside real-time physiological assessments of gastric function.

## Data Availability

All data produced in the present study are available upon reasonable request to the corresponding author.

