## Supplementary Text for "Novel Digital Gastric Alimetry® Symptom Scales for Use in Gastroduodenal Disorders and Validation against Rome IV Criteria"

**S1 - Calculations for Gastric Alimetry Symptom Scores**

Prior to calculating the four symptom scores, each symptom, with the exception of early satiety, undergoes the following transformations:

1. Overall symptom severity calculation: Continuous symptoms are averaged over time, while discrete symptoms are represented by their total count.
2. Missing Data Handling: If symptom severity data are missing, the transformed score is assigned a null value.
3. Capping: To mitigate the influence of outliers, the maximum symptom severity is limited to a predefined cap.
4. Nonlinear Transformation: An exponential scaling factor is applied to the capped scores to adjust their distribution.
5. Standardization: Linear mapping rescales the scores to a standardized range of 0 to 10.
6. Rounding: The scaled overall score is rounded to one decimal place.

The cap value and exponential scaling factor for each symptom are empirically determined based on the clinical relevance of the symptom, and aimed at achieving a near-uniform distribution of symptom severities.

The four symptom scores are then calculated by combining the relevant scaled overall scores:

| **Symptom Score** | **Contributing Symptoms** |
|---|---|
| Nausea & Vomiting Score | Nausea, Vomiting |
| Postprandial Distress Score | Early Satiation, Bloating, Excessively Full, Belching |
| Pain Score | Upper Gut Pain |
| Burning & Reflux Score | Stomach Burn, Heartburn, Reflux |

**S2 - Rules for Gastric Alimetry Symptom Tags**

Symptom tags are calculated using interpolated symptom severity curves.

| Symptom Tag | Condition |
|---|---|
| Meal Induced | (Mean of 1st hr post-meal – mean of pre-meal) > 2 |
| Meal Alleviated | (Mean of 1st hr post-meal – mean of pre-meal) < -2 |
| Late Onset | (Mean after 3rd hr post-meal – mean before 3rd hr post-meal) > 2.5 |
| Continuous | 5th percentile > 2 and (95th - 5th percentile) < 3 |

Tags are applied in the order of the rows in the above table; i.e., a symptom tagged as late onset cannot be tagged as continuous.

In addition to one of the above tags, a symptom can receive an association tag:

| Association Tag | Condition |
|---|---|
| Sensorimotor | Correlation with Gastric Amplitude ≥ 0.5 (note: correlation not calculated when variance in symptom severity is low). |

**S3 - Logistic Regression Details**

Rome IV classifications were predicted using a one-vs-rest logistic regression approach, where a separate logistic regression model was trained for each Rome IV classification. Each model estimates the probability of a patient belonging to that specific class versus all other classes.

The four Gastric Alimetry symptom scores were used as predictors. Before model training, the scores were normalized to have zero mean and unit standard deviation. The dataset was randomly split (70% training, 30% testing). The test set was used to generate Receiver Operating Characteristic (ROC) curves and evaluate model performance.

Each coefficient in the table below represents the log-odds change associated with a unit increase in the corresponding symptom score. Positive coefficients indicate that an increase in the symptom score raises the likelihood of a patient being classified into that Rome IV category. Negative coefficients indicate that an increase in the symptom score reduces the likelihood of classification into that category.

Key interpretations of the model weights are:

- CNVS classification is strongly driven by nausea and vomiting, with little influence from other symptoms.
- FD-EPS ± FD-PDS is primarily driven by pain, while nausea, vomiting, and postprandial distress scores are less relevant.
- FD-PDS only has no strong positive predictors, but a strong negative association with nausea and vomiting.

| | **CNVS +/- FD** | **FD-EPS +/- FD-PDS** | **FD-PDS only** |
|---|---|---|---|
| **Intercept** | 0.147 | -2.332 | -0.686 |
| **Nausea & Vomiting Score** | 0.447 | -0.100 | -0.433 |
| **Postprandial Distress Score** | 0.056 | -0.063 | -0.034 |
| **Pain Score** | -0.377 | 1.002 | -0.059 |
| **Burning & Reflux Score** | 0.039 | 0.003 | -0.084 |
